# Understanding Healthcare Barriers for Latino/a/e/x Families with Alzheimer’s Disease: Insights from Primary Care Provider interviews

**DOI:** 10.1101/2024.12.06.24318619

**Authors:** Diana Martinez Garcia, Maria Mora Pinzon, Jaime Perales-Puchalt

**Affiliations:** Department of Ophthalmology and Visual Sciences, School of Medicine and Public Health, University of Wisconsin – Madison, Madison, WI, USA; Department of Geriatrics and Gerontology, School of Medicine and Public Health, University of Wisconsin – Madison, Madison, WI, USA; University of Kansas Alzheimer’s Disease Center, School of Medicine, University of Kansas Medical Center, Fairway, KS, USA

**Author notes:** Corresponding author: Jaime Perales-Puchalt, University of Kansas Alzheimer’s Disease Center, Fairway, KS 66205, USA.,.

## Abstract

**Background:** Alzheimer’s Disease and Related dementias (ADRD) are disproportionately underdiagnosed, misdiagnosed, and undertreated in Latino/a/e/x populations living in the U.S. Latino/a/e/x families also experience low access to ADRD caregiver support services and high levels of depression. Primary care providers (PCPs) are the first point of contact for patients and their families, and they are critical in understanding the factors associated with disparities in accessing services. This project aims to reflect on the barriers that Latino/a/e/x families experience in accessing and using healthcare services from the perspective of PCPs.

**Methods:** The data was collected through structured interviews with 23 diverse PCPs across the US via videoconference or phone calls. Participants were recruited via snowball sampling. Two reviewers used an inductive coding approach to conduct qualitative thematic analysis. The Rigorous and Accelerated Data Reduction (RADaR) technique was used to extract relevant data and organize it into relevant categories.

**Results:** Some of the themes identified reflect the experiences of individuals in the diagnostic process and subsequent care: 1) Family members are usually the first ones to notice the symptoms, 2) Delays in seeking care might be partially influenced by denial from individuals and their families, 3) Language congruency promotes the disclosure of symptoms, 4) Care that is linguistically and literacy appropriate requires additional support of patients and families, and 5) Caregiving expectations and preferences by Latino/a/e/x families do not shield caregivers from feeling burnout. Overall, PCPs reflected that the experiences of individuals are highly influenced by socioeconomic factors, which also influence their care plans.

**Conclusion:** Most Latino/a/e/x older adults with ADRD will be cared for by a PCP at some point during their disease, which means that they require additional support and resources at primary care appointments to address the barriers to accessing care services and enhance health equity in Latino/a/e/x communities.

## INTRODUCTION

Latino/a/e/x individuals are 1.5 times more likely to develop Alzheimer’s disease and related dementias (ADRD), ^1^ have higher rates of underdiagnosis, ^2^ and have limited access to quality care.^3^ Underdiagnosis has been primarily linked to individual factors such as patient perceptions of ADRD, ^4,5^ immigration status, lack of insurance, and language preference/proficiency; ^6^ with limited research exploring how current healthcare policies contribute to the existing health disparities in diagnosis.

A 2020 study found that 80% of PCPs felt that they were on the front lines of providing care to patients with ADRD.^7^ However, PCPs often lack the training, resources, and time to educate patients and families on an ADRD diagnosis, with 70% reporting that they would be more likely to do the neurocognitive assessments with additional education.^8,9^ Because of this, patients are often referred to specialists for further testing, which results in up to half of patients not following through with those referrals.^8^

To fill the gaps in training and resources that PCPs experience, it is necessary to understand their experience in managing ADRD within the Latino/a/e/x community. This study seeks to explore those perspectives, and our results can assist in developing new programs and initiatives that provide PCPs with the tools they require to make a diagnosis in culturally appropriate and sustainable ways over time.

## METHODS

### Participant recruitment and data collection

The methodology used for recruiting participants was previously described.^10^ In summary, the PCPs were recruited using snowball sampling techniques, inclusion criteria for PCPs included being a physician (MD or DO) nurse practitioner, or physician assistant who currently or recently provided primary care services to Latine families with ADRD in the US.^10^

### Data analysis

Two reviewers independently analyzed the data inductively using qualitative content analysis.^11^ We used the Rigorous and Accelerated Data Reduction (RADaR) technique to facilitate data analysis, where the data was organized in Excel and subsequently classified according to relevance to the question and coded.^12^ The research assistant (D.M.G.) performed independent open coding of the first five transcripts and then organized them into themes, this was reviewed by the one of the authors (MMP) and discussed to ensured accuracy. Structuring the data allowed easier identification of patterns and relationships between the themes. Consistency was ensured by one of the authors (MMP), who dual-coded every fifth transcript. Once overall themes and codes were defined, the research specialist (DMG) performed another coding of the data to fit codes into an evolving collection of higher-order categories, codes were iteratively reviewed and refined to ensure that all the analytical categories were pertinent to understanding the factors influencing receiving and seeking care for Alzheimer’s Disease and Related Dementias in the Latino/a/e/x community. Once the information was summarized and organized, team members met with N.J., a PhD qualitative methodologist at the University of Wisconsin (U.W.) Institute of Clinical and Translational Research-Community Academic Partnership (ICTR-CAP) Qualitative Research Group to review the coding results and organizational details. Further modifications were made to the themes based on their feedback. Our report of this study followed the Consolidated Criteria for Reporting Qualitative Research (COREQ).^13^

### Ethics/Institutional Review Board review

The University of Kansas Medical Center Institutional Review Board approved this project (STUDY00145615). Before the interview, all participants completed an informed written consent online via their computers, tablets, or phones. A Data User Agreement between the University of Wisconsin-Madison and the University of Kansas Medical Center was executed to allow the sharing of de-identified data for this analysis. The UW-Madison Health Sciences Institutional Review Board (IRB) determined that this research met the criteria for exemption.

## RESULTS

### Participant characteristics

More than half of PCP participants were women (56.5%), US-born (52.2%), Latino (43.5%) of diverse origins, and non-Latino White (43.5%). Most PCPs were from urban settings (78.3%), with the Midwest being the most common region (65.2%). Approximately half of PCPs (52.2%) reported being able to communicate with their patients in Spanish. Most PCPs were medical doctors (65.2%) and nurse practitioners (30.4%) who reported a variation in the types of clinics they practiced in, with a wide distribution of Latino patients served.

Some quotes from participants referred to common experiences among Latino/a/e/x older adults and reflect some of the contextual factors that influence care-seeking behaviors. One of those experiences is being an immigrant who moved to the US as a senior, having emotional ties to their home country, and experiencing sadness and depression due to the continuous desire for familiar environments and social networks.

> *"Their children brought them here, people who came to the US as adults or older adults do not want to be here, they want to be home, but they cannot. If they are from [Caribbean country] and they came here when they were 60, they want to be home, so like depression, longing for home is very difficult…" (PCP 22)*

Several participants also mentioned the living arrangements and cultural practices among different generations of Latinos in the US, specifically how English language proficiency and generational status impact the level of independence seen in Latino/a/e/x individuals.

> *“… the English-speaking Hispanics that are of age, they tend to be more independent living. My first/second generation [Hispanics] it is still very much so a family unit, you have a lot of grandparents living with their kids; whereas my older Hispanics that speak English they tend to live by themselves, it is them and their wives or husbands and not so much the kids." (PCP 8)*

The final phenomena described were the experiences of US citizens compared to non-US citizens, particularly regarding the benefits or services that they might be eligible for.

> *“…the fact that we come to this country by different means affects somebody’s lifelong trajectory. Puerto Rican adults’ families have access to care, they are US citizens, so they have access to care and that is another big difference; Mexican families as well as other immigrant families… they do not always have access to care, so that I think is a difference. My patients are very well versed in their benefits, they understand how Medicare works, they know their rights, they know the benefits that they have, they still experience a lot of discrimination, so that is not different, but they for the most part can navigate those challenges whereas I think people that have a history of being an immigrant suffer disproportionately, they have not had those same opportunities.” (PCP 10)*

### Themes

The individual factors that affect how care is provided and received are explored according to when they occur in the care continuum: 1) Recognition of symptoms and care-seeking behaviors, 2) Interacting with the healthcare system to get a diagnosis and subsequent care, and 3) Experiencing the disease-process as a family.

#### Recognition of Symptoms & Care Seeking Behaviors (Table 1)

##### Family members are usually the first ones to notice symptoms

In the experience of healthcare providers, many times, a family member expressed concern for the patient’s health, which first led to the exploration of further diagnosis. Providers required patients to have informants who lived with them to better understand how advanced the cognitive decline was and what additional resources were needed. Familiarity with the patients and their families was also identified as important in identifying symptoms and being able to compare and contrast symptoms according to patients and their informants.

**Table 1:**
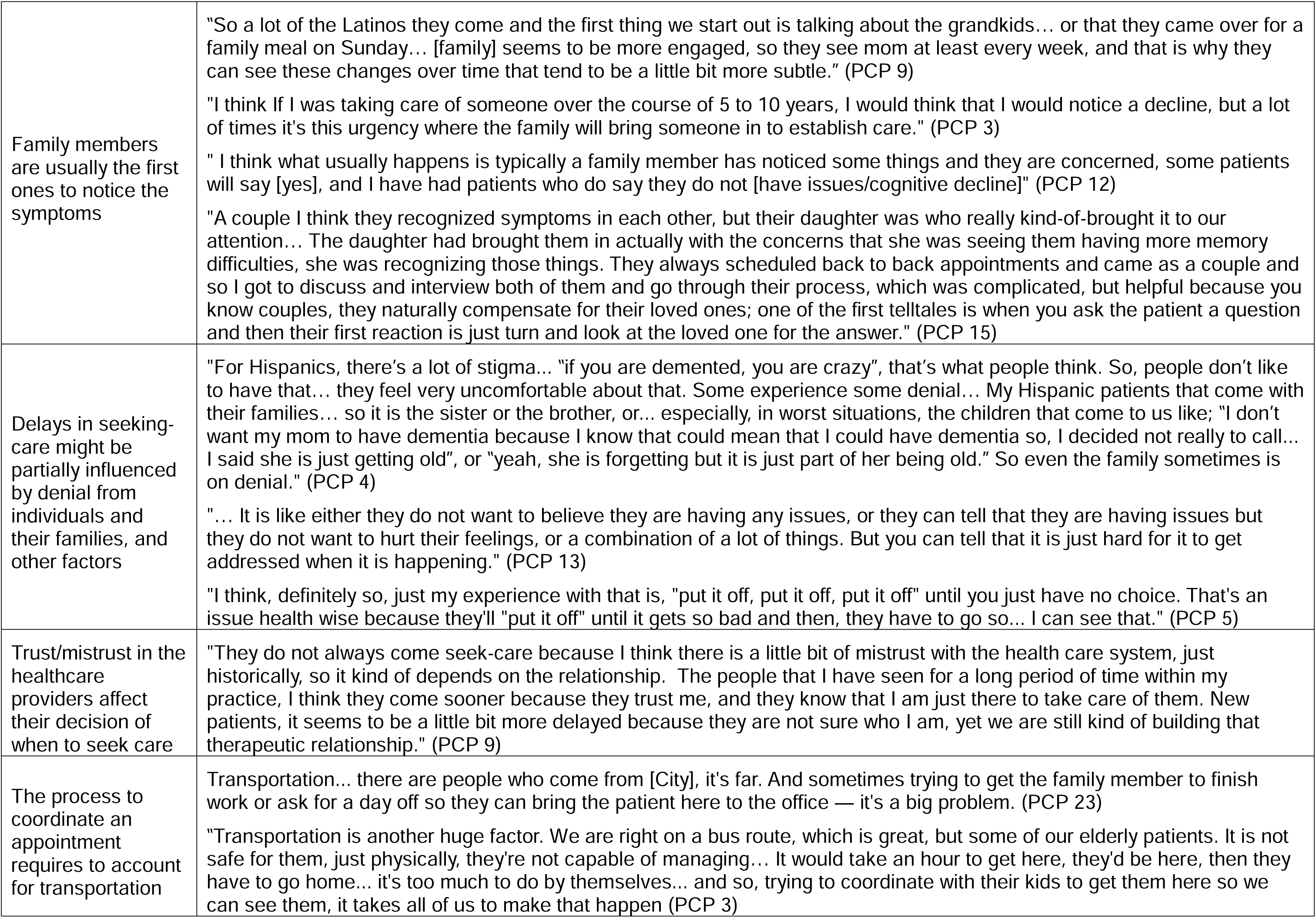
Quotes reflecting on the Recognition of Symptoms & Care Seeking Behaviors.

|  |  |
| --- | --- |
| Family members are usually the first ones to notice the symptoms | "So a lot of the Latinos they come and the first thing we start out is talking about the grandkids... or that they came over for a family meal on Sunday... [family] seems to be more engaged, so they see mom at least every week, and that is why they can see these changes over time that tend to be a little bit more subtle." (PCP 9)<br>"I think If I was taking care of someone over the course of 5 to 10 years, I would think that I would notice a decline, but a lot of times it's this urgency where the family will bring someone in to establish care." (PCP 3)<br>" I think what usually happens is typically a family member has noticed some things and they are concerned, some patients will say [yes], and I have had patients who do say they do not [have issues/cognitive decline]" (PCP 12)<br>"A couple I think they recognized symptoms in each other, but their daughter was who really kind-of-brought it to our attention... The daughter had brought them in actually with the concerns that she was seeing them having more memory difficulties, she was recognizing those things. They always scheduled back to back appointments and came as a couple and so I got to discuss and interview both of them and go through their process, which was complicated, but helpful because you know couples, they naturally compensate for their loved ones; one of the first telltales is when you ask the patient a question and then their first reaction is just turn and look at the loved one for the answer." (PCP 15) |
| Delays in seeking-care might be partially influenced by denial from individuals and their families, and other factors | "For Hispanics, there's a lot of stigma... "if you are demented, you are crazy", that's what people think. So, people don't like to have that... they feel very uncomfortable about that. Some experience some denial... My Hispanic patients that come with their families... so it is the sister or the brother, or... especially, in worst situations, the children that come to us like; "I don't want my mom to have dementia because I know that could mean that I could have dementia so, I decided not really to call... I said she is just getting old", or "yeah, she is forgetting but it is just part of her being old." So even the family sometimes is on denial." (PCP 4)<br>"... It is like either they do not want to believe they are having any issues, or they can tell that they are having issues but they do not want to hurt their feelings, or a combination of a lot of things. But you can tell that it is just hard for it to get addressed when it is happening." (PCP 13)<br>"I think, definitely so, just my experience with that is, "put it off, put it off, put it off" until you just have no choice. That's an issue health wise because they'll "put it off" until it gets so bad and then, they have to go so... I can see that." (PCP 5) |
| Trust/mistrust in the healthcare providers affect their decision of when to seek care | "They do not always come seek-care because I think there is a little bit of mistrust with the health care system, just historically, so it kind of depends on the relationship. The people that I have seen for a long period of time within my practice, I think they come sooner because they trust me, and they know that I am just there to take care of them. New patients, it seems to be a little bit more delayed because they are not sure who I am, yet we are still kind of building that therapeutic relationship." (PCP 9) |
| The process to coordinate an appointment requires to account for transportation | Transportation... there are people who come from [City], it's far. And sometimes trying to get the family member to finish work or ask for a day off so they can bring the patient here to the office — it's a big problem. (PCP 23)<br>"Transportation is another huge factor. We are right on a bus route, which is great, but some of our elderly patients. It is not safe for them, just physically, they're not capable of managing... It would take an hour to get here, they'd be here, then they have to go home... it's too much to do by themselves... and so, trying to coordinate with their kids to get them here so we can see them, it takes all of us to make that happen (PCP 3) |

##### Delays in seeking care might be partially influenced by denial from individuals and their families and other factors

Latino/a/e/x patients tended to wait until severe symptoms presented to seek care for themselves, while family members of those with a diagnosis of dementia did not seek care for the patient until they felt they could not take care of their loved one appropriately. Symptoms are brushed off as "a normal part of aging" until the cognitive decline of their loved one is too evident. This delay in seeking care was attributed to fear of their loved one being labeled as “crazy” and fear of themselves developing ADRD in old age.

##### Trust/mistrust in healthcare providers affects their decision when to seek care

In the Latino/a/e/x community, mistrust in the healthcare system might be due to fear of deportation and implications on benefits or immigration procedures, among others. Due to this mistrust, patients might hesitate to seek medical care, which delays necessary treatment and impacts their overall health outcomes. Trust-based relationships with healthcare providers can be built by providing culturally sensitive care and listening to patient concerns.

##### The process of coordinating an appointment requires accounting for transportation

One of the main barriers to treating Latino/a/e/x patients was the distance to be seen and the need for transportation, as many older adults do not drive and rely on public transportation or rides from family or friends. Depending on the advancement of the disease, there are times when patients cannot travel alone or when an informant is needed, so appointments need to account for work schedules for care partners.

#### Interacting with the healthcare system to get a diagnosis and subsequent care (Table 2)

##### Accurate diagnosis requires further assessments that might not be available due to the patient’s insurance (or lack thereof)

Healthcare providers often employ varied diagnostic methods and treatment options, which not only reflect their unique backgrounds and level of comfort with managing ADRD but also consider the type of insurance (or lack thereof) a patient possesses. It highlights the financial barriers involved in referring patients for specialized care. For instance, patients are offered low-cost initial visits to ensure access to affordable medical care. However, the cost can become a barrier when specialized care is required. Overall, it reflects the complexity of providing comprehensive care where financial challenges are significant.

**Table 2:** Quotes reflecting on the interactions with the healthcare system to get a diagnosis and subsequent care.

|  |  |
| --- | --- |
| Language congruency promotes the disclosure of symptoms. | "Many of them, of course, are surprised that they have a doctor who can speak their language, and then they open up a little more, they feel more comfortable being able to express their complaints or their symptoms. Explain a little better. And I receive them almost every day when I see patients who speak Spanish they say, "doctor, how wonderful that I can explain everything to you... and how I feel", so that satisfies them, of course it also improves communication. with the patient." (PCP 7)<br>"Sometimes it just takes longer because they just feel more comfortable giving you much more details, and when the nurse collects because once they don't need to use the interpreter is like, "Oh, I have knee pain but is not actually the knee, it's actually that my back hurts and now I don't feel... I am not able to walk and it's actually my jaw and I have this callus here, and I have back pain and all of these things." (PCP 4) |
| Accurate diagnosis requires further assessments that might not be available due to the patient's insurance (or lack thereof) | "Here, we charge \$15 a visit and we never turn anybody away if they can't pay... But the real struggle is what happens when we want an external referral. We would really need like a neurologist... we also might need to do labs; there's so much medical workup that we can do. Well, at [Medical Center]... we would have to work with the financial assistance department... They [community Health Worker] might help the family fill out the paperwork, so that we could get the care without a financial burden. A lot of logistics has to happen. That's why, in a way, it could be burdensome to make that referral. These things kind of happen incrementally and it depends on really what the family is capable of partnering with us to do. (PCP 3)<br>"They [patient] could not understand why I could not fix it, or why I cannot address it, and I just was not in a place where I felt comfortable starting people on medications without a more thorough exam... if people are uninsured, they definitely got charged less, but still an MRI is really expensive, laboratory work is very expensive. So it is often like trying to negotiate with them on what they are willing to do and not do, and they did not really want to get seen or they would be wanting to get seen. (PCP 13) |
| Care that is linguistically and literacy appropriate requires additional support of patients | "If I know that they are illiterate, I try to create extra ways to help them, for example... create certain schedules. I am very visual so; I have no problem with just drawing things for my patients. It takes extra time so, that's why I am telling that very often it is just time consuming when I have a new patient who feels that I am the person who they can talk about everything... I have referred them to some websites, and whatever thing I have found for them in Spanish. I do have a large list of handouts that I have access to... not just like for dementia but, for other medical problems like hypertension, diabetes, whatever." (PCP 4)<br>"A lot of the family members, go to those appointments and they do not know the medical terminology, so even though if they have their family member or interpreter what they actually got from the visit is not what they should have got from the visit, and once I get the consultant's letter we go over what they said, but sometimes they do not even understand what they were told. We do have a lot of that when patients come back and they are not on their medication and their treatment sounds like okay, let us go back and read their note and see what they said and what they wanted, that is a challenge too." (PCP 11)<br>"And sometimes when they talk to the neurologist on the phone... people sometimes don't answer the phone if they don't know what number it is. There are people who do not answer it, even if there are immigration problems, if they do not know what the number is, they are not going to answer because they are afraid that they are going to be deported or something like that. So yes, sometimes there is that barrier to establishing that connection with the specialist who is the neurologist." (PCP 7) |

##### Language congruency promotes the disclosure of symptoms

Most providers could access translation services at their institution if they did not speak Spanish. These services were provided free of charge according to policy and regulations. However, the methods of accessing them varied widely (e.g., in person, over the phone, video conferencing), and each involved different barriers. The two most common barriers to using translation services were the coordination required to access the translator and the extra time their use added to the visit. Over-the-phone translation services were identified as not ideal for evaluating cognitive decline as it creates confusion. When congruent language between patients and providers was available, it increased patient comfort.

##### Care that is linguistically and literacy appropriate requires additional support for patients

Providers expressed their patients’ interest in receiving linguistically and literacy-appropriate care, which sometimes requires drawing and creating resources from scratch to match the educational history and native language of patients. Furthermore, instructions about follow-up appointments and the next steps in the process should be considered, as patients might not answer the phone to unknown numbers.

#### Experiencing the disease process as a family (Table 3)

##### Caregiving expectations and preferences do not shield caregivers from feeling burnout

Strong family-oriented values and a sense of responsibility are heavily influential within the Latino/a/e/x community. There is a preference to care for family members rather than placing them in an assisted living facility after their diagnosis. Given the stigma associated with living facilities within the Latino/a/e/x community, caring for their loved ones at home ensures the best care. However, caregivers also work and often experience stress and exhaustion, which might push them to seek support from their loved ones’ providers to find ways to overcome their overwhelming responsibilities.

**Table 3:** Quotes reflecting on experiencing the disease process as a family.

|  |  |
| --- | --- |
| Caregiving expectations and preferences do not shield caregivers from feeling burnout | • “There has been a couple of times where we have had individuals either come back into the office, or put their significant other in the car and come back in, and ask to talk because they are tired or stressed. Because they are still having to work along with taking care of them.” (PCP 17)<br>• “Individuals do not want to put their family member in a home... especially in the Latino population, they really are strong family people, they really love their family members, and they would do anything for them and they really like to do that for themselves and not anyone else do it for them because they want to make sure that they are well taken care of.” (PCP 17)<br>• “They are multigenerational home, so the daughter has teenagers and now mom moved in and now they are trying to take care of that... but there is more spousal support overall. I had a son-in-law bring in their mother to clinic visits and that is just not common throughout every culture. So yeah, definitely caregiver stress, they manage a lot of things at once usually. “ (PCP 19)<br>• "I think a lot of times those patients are worried if they can stay in the home and not put them in a [nursing] home, I think that is worry amongst all individuals usually because there is that stigma about those places unfortunately, so I think that is a main concern and then obviously having the resources specifically for them and making sure that they were right for them I guess the right people to go to and what not." (PCP 17) |

## DISCUSSION

We describe the challenges PCPs observe in Latino/a/e/x patients seeking an ADRD diagnosis and the potential ways to reduce them. Understanding the challenges Latino/a/e/x families face through the insights of PCPs is crucial for developing effective, equitable, and culturally appropriate healthcare strategies for ADRD. This perspective is important because PCPs are often the first point of contact for most Latina/e/x families with ADRD,^14^ and their role in ADRD care is expected to increase even more to reduce long waits in delivering anti-amyloid medications.^15^ Effective interventions can empower the Latino/a/e/x community to take an active role in seeking care or an ADRD diagnosis and can lead to the development of support systems for caregivers.

Consistent with prior research, our findings underscore the critical role of family in the early detection of ADRD symptoms.^16^ Because Latinos are more likely to be in multigenerational homes,^17^ or have close family connections, this ultimately allows the family members to be the first people to recognize abnormal changes in their loved ones’ behavior. However, our study also highlights that families can contribute to delays in seeking care, as families need to coordinate schedules across multiple members, might overlook symptoms, or when concerns related to immigration or legal issues lead to postponed care, including in those who being US citizens, live in a mixed-status household.^18^ Our study also identifies another cause for delays, which is the fear associated with developing dementia if a parent already has it as one of the contributing factors. Overall, regardless of the cause, delays exacerbate disease progression, complicating management, which supports the need for culturally sensitive educational interventions to mitigate such delays and promote a better engagement of families in the healthcare process.^19,20^ Asking families and their older patients about their loved one’s cognitive status may also prompt the use of screening and reduce those delays,^21^ as this would leverage the family’s awareness of decline and the PCP’s clinical expertise.

Our study also reaffirms the centrality of *’familismo’* in the health behavior of the Latino/a/e/x community, with multigenerational homes facilitating early symptom recognition.^22^ Despite a trend towards individualism in second or third-generation U.S.-born Latino/a/e/x individuals, our study found a consistent preference for family members as caregivers across generations to ensure that the person with dementia receives the best treatment, suggesting a persistent cultural influence that withstands acculturation pressures. This agrees with previous research, which found that Latino/a/e/x individuals do not question the need to provide care to their family members.^23^ Nonetheless, our study adds to the discourse by emphasizing the reality of caregiver burnout, a concern often overshadowed by the cultural expectation of familial caregiving.^16^ This highlights a discrepancy between cultural values and practical caregiving challenges, necessitating a reevaluation of support systems for caregivers.

Latino/a/e/x families with a strong social network report extensive support with the division of responsibilities to care for and help manage the stress that an ADRD diagnosis creates. However, not all patients have multiple family members taking care of them; thus, their caregivers experience a significant adjustment to their lives. These caregivers have expressed feelings, at times, of being trapped in their role and are often frustrated by the limited family support. As the health of the patient with ADRD deteriorates, caregivers are tasked with managing the increasing responsibilities and very frequently express concern for both their and their family members’ future, as some feel ill-prepared for what the progression of the disease may look like.

Our study corroborates the literature that shows that language congruency between patients and PCPs is pivotal for effective care.^24^ Language congruence promotes the disclosure of symptoms and experiences that might otherwise get lost in translation and allows for better care overall. In a study done in early 2023, all 18 participants indicated they felt they received better care from their provider due to them speaking the same first language during their appointments. Additionally, there is a relationship between language-concordant care and stronger patient-provider trust, ultimately allowing patients to feel more comfortable and satisfied with the care they receive than having an interpreter.^24^

Our study also highlights the logistical challenges and extended visit durations introduced by the use of translation services when language congruency is unavailable. Providers noted that phone interpreter services cannot read non-verbal queues, which are often present in cognitive decline and thus allow for in-person and virtual translator options to provide a better experience for patients.^25^ The main barrier to using the translation services was the required extra time needed for appointments, which was rarely accounted for, resulting in providers having to adjust their schedules or affect the quality of care.

Our results echo the literature’s identification of socioeconomic factors as significant determinants of healthcare access for Latino/a/e/x individuals with ADRD.^26^ Our study further highlights how health centers and healthcare providers navigate patients’ ability to pay, insurance coverage, and geographical barriers that sustain health inequities. While diagnostic methods and treatment options varied, services were provided based on a sliding scale fee, and some clinics offered discounted or free medications. However, the common factor was that healthcare facilities had to create an exhaustive patient-based process that allowed these social factors to be considered and accommodated in their appointments.

The study acknowledges limitations, including a small subset of PCP perspectives and a focus on urban settings in the Midwest region of the United States, where Hispanic/Latino populations are a minority, which might limit the generalizability of results to other locations with more experience and resources to assist Latino communities. Future research should expand the geographical scope and consider the specific resources available to the participants’ patient population to enhance the generalizability of findings.

In conclusion, our study calls for targeted efforts to support Latino/a/e/x families in the ADRD care continuum. It highlights the need for culturally competent care plans and resources, addressing the intricate interplay of family dynamics, cultural values, language and literacy needs, and socioeconomic constraints. By doing so, healthcare providers can better serve this vulnerable population and reduce disparities in ADRD diagnosis, treatment, and support. The discussion integrates these themes within the broader context of existing academic discourse, providing a foundation for future research and policy development in this critical area of healthcare.

## Data Availability

All data produced in the present study are available upon reasonable request to the authors.

## Acknowledgments

Dr. Perales-Puchalt thanks the national and local organizations that have partnered with him to conduct present and past research since 2015. The research team thanks research participants included in all stages of this research as well as anyone who has contributed directly and indirectly to this research. The ideas and opinions expressed herein are those of the authors alone, and endorsement by the authors’ institutions or the funding agency is not intended and should not be inferred.

## Disclosure of Use of AI

During the preparation of this work the author(s) used Grammarly, and Microsoft Copilot to proofread and improve readability of the introduction and discussion. After using these tools, the author(s) reviewed and edited the content as needed and take(s) full responsibility for the content of the publication.

